# Increased blood–brain barrier leakage in schizophrenia spectrum disorders compared to healthy controls in dynamic contrast-enhanced magnetic resonance imaging

**DOI:** 10.1101/2023.12.12.23299782

**Authors:** Joanna Moussiopoulou, Vladislav Yakimov, Boris-Stephan Rauchmann, Hannah Toth, Julian Melcher, Iris Jäger, Isabel Lutz, Marcel Kallweit, Boris Papazov, Klaus Seelos, Amir Dehsarvi, Lukas Röll, Mattia Campana, Florian Raabe, Isabel Maurus, Peter Falkai, Alkomiet Hasan, Nicolai Franzmeier, Daniel Keeser, Elias Wagner

## Abstract

**Background:** There is growing evidence for inflammatory mechanisms in schizophrenia spectrum disorders (SSD) that have been associated with blood-brain barrier (BBB) disruption. Previous studies investigating the BBB in SSD focused on cerebrospinal fluid (CSF) markers, that cannot adequately assess BBB integrity. Dynamic contrast-enhanced magnetic resonance imaging (DCE-MRI) represents a sensitive method for investigating subtle barrier breakdown in vivo. So far, only one pilot study has investigated BBB breakdown in SSD with DCE-MRI, in a relatively small cohort. We hypothesized higher leakage in SSD compared to HC, indicative of a clinical sub-phenotype of SSD.

**Methods:** Forty-one people with SSD and 40 age- and sex-matched healthy controls (HC) were included in the final analyses of the cross-sectional study. DCE-MRI, clinical characterization, cognitive assessments, blood and CSF analyses were conducted. The volume transfer constant K_trans_ was calculated with pharmacokinetic modelling (Patlak method), to estimate the rate of contrast agent transfer between blood and the brain’s extravascular space. K_trans_ maps were compared between the groups to detect group differences in BBB leakage. Within the SSD cohort, the association between leakage and clinical characteristics was investigated with linear regression analyses.

**Results:** Group comparisons of K_trans_ maps showed higher leakage in SSD compared to HC on a whole brain level. The effect was more pronounced in first episode compared to multiple episode psychosis. No association was detected between leakage and measures of cognition, psychopathology, peripheral inflammation and albumin CSF/serum ratio.

**Discussion:** This is the largest study to date investigating the BBB in SSD with DCE-MRI in a multimodal approach, allowing direct exploration of the BBB, compared to a HC group. The integrity of the BBB is crucial for maintaining the brain’s microenvironment, and its disruption could be associated with potential immune system abnormalities. The results of this study provide the first in vivo evidence of higher BBB leakage on a whole brain level compared to HC. The disruption of the BBB in SSD, as detected through DCE-MRI, may provide insights into the disease’s mechanisms and potential for targeted treatments. Further research in this area may clarify specific biological disease mechanisms and identify new therapeutic targets.

## Introduction

Schizophrenia spectrum disorders (SSD) are a multidimensional and multifactorial disease entity characterized by positive (delusions, hallucinations), negative (social withdrawal, apathy), and cognitive symptoms (poor executive function and memory. Even before disease onset, general cognitive performance is on average 1.5 standard deviations below that of the general population [1] and cognitive impairment determines the degree of disability [2]. Mounting evidence indicates the contribution of immune-mediated / neuroinflammatory factors [3, 4], first postulated over a century ago and further supported by more recent epidemiological, biomarker, neuroimaging, post-mortem, interventional and genome-wide association (GWAS) studies [5, 6].

Blood-brain barrier (BBB) breakdown due to changes on ultrastructural level (e.g. disruption of tight junctions), and inflammatory processes may initiate and/or contribute to progressive synaptic and neuronal dysfunction and neurodegenerative disorders [7]. BBB breakdown is presumed to be an early event in the aging human brain that begins in the hippocampus and may contribute to cognitive impairment [8].

The BBB is a highly selective semipermeable structural and functional barrier that is anatomically comprised by endothelial cells (in addition to astrocytes and pericytes), interconnected by tight junctions, which prevents peripheral neurotoxins and pathogens from entering the brain [9–11]. Besides small molecules like oxygen that can cross the BBB [12], the remainder is subject to specialized transport systems in order to pass from blood plasma to brain tissue or to be removed from the brain [13]. The blood-cerebrospinal fluid barrier (BCSFB) is comprised by the epithelial cell monolayer of the choroid plexus (ChP), surrounding a complex vascular convolute of fenestrated capillaries and connective tissue, located in the lateral, third and fourth ventricles [14]. Maintaining a structurally and functionally intact BBB and BCSFB is pivotal to maintain homeostasis of the CNS, a fundamental prerequisite for neuropsychiatric well-being.

Various CNS diseases such as stroke [15], Alzheimeŕs disease (AD) [16] and multiple sclerosis (MS) [17] have been associated with a loss of BBB integrity, resulting in leakage of blood components into the CNS, disrupted clearance of molecules and cellular infiltration [9, 10]. BBB breakdown is associated with inflammatory mechanisms (i.e. upregulation of inflammatory cytokines and reactive oxygen species) [18] and is presumed to be an early process in the pathological cascade leading to neurodegeneration [8, 19]. Neurodegenerative processes or processes of failed neuro-regeneration develop at least partially because of inflammatory responses [20]. The inflammation resulting from autoimmune or infectious diseases might increase the permeability of the BBB, thus exposing CNS tissues to molecules such as cytokines or antibodies and hence possibly increasing the chance of an activation of resident inflammatory cells and infiltration of innate immune cells [21]. This vicious cycle of neuroinflammation and BBB disruption has been already reported elsewhere [3, 22]. To date, a first scientific evidence base is emerging demonstrating that routine CSF analyses in individuals with psychosis reveal markers of inflammatory or infectious aetiology [23], in addition to the detection of autoimmune encephalitis or autoimmune psychosis [24]. The complex nature of BBB dysfunction in psychosis has been linked with disrupted neuronal and synaptic function, increased permeability to inflammatory molecules, disrupted glutamate homoeostasis, impaired action of antipsychotics, development of antipsychotic resistance [25] and is reported to play a key role in the genesis of neurovascular dysfunction and associated neurodegeneration [26]. All these aspects can in turn possibly contribute to cognitive and behavioral symptoms in SSD.

The BBB can currently be estimated through post-mortem, serum and cerebrospinal fluid (CSF)- biomarker, and neuroimaging studies. Elevated albumin (Alb) in the (lumbar) CSF has numerously been described to suggest an opening in the BBB, making the CSF/blood Alb ratio (Q_Alb_) a clinically broadly used (gold standard) biomarker for BBB breakdown [25, 27]. Of note, the status of Q_Alb_ in BBB evaluation is questionable due to multiple reasons. Firstly, its implementation is challenged due to its invasive method of obtainment (lumbar puncture). Furthermore, Q_Alb_ can be elevated due to reasons other than barrier breakdown [25]. In addition, it predominantly reflects the status of the *BCSFB*, rather than the BBB itself [28], due to its circulation direction [29] (**suppl. figure S1**). The prevalence of BCSFB breakdown in SSD (estimated through Q_Alb_) has been reported around 20% [25]. The implication of both the blood-brain and the blood-cerebrospinal fluid barriers and in the etiology and clinical trajectories in SSD remains elusive. In summary, currently available modalities only enable indirect estimations of the BBB or investigate the status of the *BCSFB* rather than the BBB. Bridging the gap between the periphery and the brain remains a challenge and a limited understanding of the BBB has hindered its direct targeting in disease and therapy [26].

Among in-vivo methods, dynamic contrast-enhanced (DCE) MRI stands out as a minimally invasive tool to detect BBB breakdown. Unlike approaches such as Q_Alb_, DCE-MRI offers a specific assessment of BBB integrity, allowing direct conclusions about its status, while also enabling localization of affected brain areas, with high spatial and temporal resolution [8]. It is based on the extravasation (leakage) of intravenously injected contrast agent (CA) into the extravascular space, which results in image contrast enhancement that can be quantified. The efflux rate of the CA from plasma into the brain tissue can be calculated through pharmacokinetic modelling of DCE T1-weighted MRI signal intensities, from which different permeability measures can be quantified [15, 30]. It is the method of choice in permeability imaging [31] with moderate-to-excellent reproducibility [32], but has not been implemented in SSD research so far. Moreover, it has been proven to be the most advanced method of investigating *subtle* (when compared to intensive) leakages [33] [34], which makes it particularly favorable in BBB investigations in SSD, a challenge given the low permeability values being considered in this cohort [35, 36]. DCE-MRI combined with kinetic modelling (i.e. the Patlak model) has been described as the most accurate and appropriate way to investigate low expected leakage [8, 37]. Of note, estimations of the volume transfer constant K_trans_ have been found to be robust to the model assumptions [34].

To our knowledge only one study so far has investigated the BBB integrity in SSD, with DCE-MRI, in a limited sample size. Cheng et al. reported higher K^trans^ values of the bilateral thalamus in a schizophrenia (SCZ) group compared to a healthy control (HC) group (N= 29 SCZ vs. 18 HC), but these findings render further replication.

The present study is the first to investigate BBB breakdown and leakage in a large SSD cohort, compared to age and sex-matched healthy control individuals (HC) (total N= 81) using DCE-MRI. We hypothesized that due to the frequently postulated CNS barrier disruption in SSD, there would be a significant higher leakage in SSD compared to HC (**hypothesis 1 – main hypothesis**). Furthermore, we postulated, that the higher leakage reflects a clinical sub-phenotype of SSD (**hypothesis 2**), and that higher leakage correlates positively with systemic inflammatory processes (**hypothesis 3**) and BCSFB breakdown (**hypothesis 4**).

## Methods

### Participants

A total of 45 people with SSD treated at the Department for psychiatry and psychotherapy of the LMU University Hospital, Munich, Germany, were enrolled between March 2022 and October 2023. Inclusion criteria comprised age between 18 and 60 and a diagnosis of SSD, according to DSM-V, assessed with the Mini International Neuropsychiatric Interview (M.I.N.I.), German version 7.0.2 [38].

In addition, 42 age- and sex-matched healthy controls (HC) were recruited via announcements in digital channels (e.g. homepage of the university hospital, social media) and enrolled between June 2022 and October 2023. HC were defined as participants with no past or current self-reported psychiatric disorder. HC were also aged between 18 and 60 years. Exclusion criteria for both groups were defined as follows: a primary psychiatric disorder other than SSD (only SSD cohort), any central nervous system (CNS) disorder, severe internal medicine, inflammatory, rheumatic diseases, current pregnancy or lactation, regular current drug abuse (in the past month), coercive treatment, acute suicidality, inability to give informed consent, current participation in clinical trials, current electroconvulsive therapy or other brain stimulating therapies, and contraindication(s) to MRI or DCE-MRI, such as renal failure. Prior to inclusion in the study, all participants provided written informed consent. The study protocol was approved by the ethics committee the Ludwig-Maximilian University Munich (reference number 21-1139).

### CSF and blood acquisition

All individuals with SSD underwent basic blood test including complete blood count analysis as part of routine clinical care. Neutrophil-to-lymphocyte ratio (NLR), monocyte-to-lymphocyte ratio (MLR) (ratios from the absolute counts) were selected as indicative for inflammatory and immune-related processes [39]. Where clinically indicated in the context of exclusion diagnostics, CSF examinations were offered and conducted, as recommended by the German evidence- and consensus-based schizophrenia guidelines [40]. Q_Alb_ was selected as indicative for CSF pathology or BCSFB disruption and adjusted by age according to the formula: Q_Alb_ = (4+age/15) × 10−3 [41], age indicated as years. In all cases, the CSF and serum were tested for neuronal autoantibodies (**suppl. methods**) to rule out autoimmune-mediated encephalitis.

### Demographic, clinical, and cognitive assessments

All individuals underwent basic demographic and clinical assessments (BMI, total education, medication intake, somatic conditions, substance abuse, current smoking status), collected via interviews and questionnaires and, if possible, verified using medical records. In individuals with SSD the following additional variables were collected through self-report and verified on medical reports whenever possible: information on current antipsychotic medication (current antipsychotic medication were converted to chlorpromazine equivalent doses (CPZequivalent) [42]), lifetime antipsychotic medication in months, duration of illness (DUI, defined as duration since first being diagnosed), first episode (FEP) or multiple episode (MEP) status (FEP being defined as first hospitalization due to psychotic symptoms). Cognitive performance was assessed with the Trail Making Test (TMT, part A and B) in both groups. In Part A, participants were asked to link numbered points randomly distributed on a sheet of paper in ascending order as rapidly as possible, without removing the tip of the pen from the page. In Part B, the participants were asked to link numbers and letters alternately. The time of test performance was measured by a trained researcher. TMT accesses the cognitive elements attention, visual search and scanning, processing speed, task switching, cognitive flexibility and executive function [43]. Patients additionally underwent the Mini International Neuropsychiatric Interview (M.I.N.I.) interview for diagnosis validation [38], the symptom severity assessment using the Positive and Negative Syndrome Scale (PANSS) [44], and the Global Assessment of Functioning (GAF) Scale [45].

### Structural and DCE-MRI acquisition

MRI images were acquired on a 3-Tesla MRI scanner (3T Magnetom Prisma, Siemens Healthcare GmbH) in the Department of Psychiatry and Psychotherapy, University Hospital, LMU Munich, Munich, Germany, with a 64-channel head coil. **suppl. figure S2** illustrates the data acquisition and analysis steps, **suppl. Table 1** shows technical details of the scanning methodology.

The imaging protocol included structural sequences for anatomical reference and clinical evaluation (T1-weighted magnetization-prepared rapid gradient-echo (MP-RAGE) and T2-weighted sampling perfection with application optimized contrast using different flip angle evolution (SPACE) sequences). The dynamic contrast–enhanced (DCE) sequence followed for leakage calculations. This sequence consisted of 23 minutes total scan time with a voxel size of 1.8 × 1.8 × 1.8 mm. During this sequence (after approximately 2 minutes, scanning time point 12/80), the gadolinium-based contrast agent (GBCA) (Gadobutrol, Gadovist®, Bayer AG, Leverkusen, Germany, 0.1 mmol/kg) was injected in the antecubital vein (injection rate 3 mL/s, followed by 25-30 mL saline flush). All scans were evaluated clinically by an experienced neuroradiologist, and pathological scans were excluded from the analyses.

### Data processing and analysis Statistical analysis of demographics

The following tests were used to compare demographic characteristics between SSD and HC groups and clinical characteristics within the SSD cohort, with Rstudio, version 4.1.2, 2021 [46]. Group differences in sample characteristics were explored with Chi-squared test for categorical variables, Welch’s two sample t-test for normally distributed, and Mann–Whitney U test for non-normally distributed continuous variables. Normality within groups was assessed using the Shapiro-Wilk test. The threshold for statistical significance was set at p-value < 0.05. Descriptive statistics are shown as mean ± standard deviation (**Tables 1-3**).

**Table 1.** Demographic data of the individuals that were included in the DCE MRI analyses. Y= yes, n=no, BMI= Body Mass Index, TMT= Trail Making Test, W= Wilcoxon rank-sum, SD= standard deviation, df= degrees of freedom

|  |  | SSD |  | HC |  | SSD vs. HC |  |  |
| --- | --- | --- | --- | --- | --- | --- | --- | --- |
| N (SSD/HC) |  | 41 |  | 40 |  | Chi2 | df | p |
| **Demographics** |  |  |  |  |  |  |  |  |
| Sex (f : m) | 81 (41/40) | 10:31 |  | 11:29 |  | 0.004 | 1 | 0.95 |
| Current smoking (y : n) | 81 (41/40) | 22:18 |  | 5:35 |  | 14.31 | 1 | **0.00015** |
|  |  | **Mean** | **SD** | **Mean** | **SD** | **t/W\*** | **df** | **p** |
| Age (yrs) | 81 (41/40) | 35.17 | 9.77 | 33.55 | 9.60 | 897* | - | 0.47 |
| Total education (yrs) | 77 (37/40) | 14.84 | 3.59 | 18.39 | 2.14 | - | 57.73 | **2.57e-06** |
| BMI (kg/m<sup>2</sup>) | 80 (40/40) | 27.14 | 4.50 | 24.21 | 4.45 | 1110* | - | **0.0029** |
| **Cognition** |  |  |  |  |  |  |  |  |
| TMT A | 72 (32/40) | 33.31 | 13.67 | 23.10 | 6.36 | 933* | - | **0.0009** |
| TMT B | 72 (32/40) | 92.78 | 64.34 | 54.55 | 19.02 | 954.5* | - | **0.0004** |

**Table 3.** Demographic data of the SSD cohort according to their FEP/MEP status. Y= yes, n=no, TRS= treatment resistance, assessed through lifetime Clozapine intake, lifetime AP= lifetime antipsychotic treatment, CPZ= Chlorpromazine, GAF= Global Assessment of Functioning Scale, PANSS= Positive And Negative Syndrome Scale, pos= positive, neg=negative, gen= general, tot= total, TMT= Trail Making Test, W= Wilcoxon rank-sum, SD= standard deviation, df= degrees of freedom

|  |  | FEP |  | MEP |  | FEP vs. MEP |  |  |
| --- | --- | --- | --- | --- | --- | --- | --- | --- |
| N (FEP/MEP) |  | 11 |  | 30 |  |  |  |  |
|  |  |  |  |  |  | Chi2 | df | p |
| Demographics and Disease characteristics |  |  |  |  |  |  |  |  |
| Sex (f : m) | 41 (11/30) | 1:10 |  | 9:21 |  | 0.94 | 1 | 0.33 |
| Current smoking (y : n) | 41 (11/30) | 7:4 |  | 16:14 |  | 0.05 | 1 | 0.82 |
| TRS (y : n) | 41 (11/30) | 0:11 |  | 15:15 |  | 6.65 | 1 | **0.01** |
| Andreasen remission criteria (y : n) | 37 (9/28) | 1:8 |  | 5:23 |  | 1.95e-32 | 1 | 1 |
|  |  | Mean | SD | Mean | SD | t/W* | df | p |
| Age (yrs) | 41 (11:30) | 33.64 | 10.35 | 35.7 | 9.68 | 187* | - | 0.53 |
| Total education (yrs) | 37 (9/28) | 14.39 | 2.40 | 14.98 | 3.93 | 0.54 | 22.76 | 0.59 |
| BMI (kg/m²) | 40 (11/30) | 26.43 | 3.76 | 27.38 | 4.76 | 0.64 | 19.42 | 0.53 |
| Duration of illness (DUI) (months) | 41 (11/30) | 7.59 | 8.45 | 147.5 | 114.34 | 327* | - | **1.96e-06** |
| CPZ equivalent [mg] | 41 (11/30) | 203.48 | 111.33 | 481.40 | 275.49 | 274.5* | - | **0.001** |
| Lifetime AP (months) | 41 (11/30) | 1.30 | 1.23 | 119.20 | 113.43 | 313.5* | - | **1.323e-05** |
| GAF | 41 (11/30) | 51.91 | 8.72 | 49 | 13.68 | 131* | - | 0.32 |
| Psychopathology |  |  |  |  |  |  |  |  |
| PANSS total | 38 (9/29) | 63.78 | 13.60 | 62.07 | 13.03 | -0.33 | 12.91 | 0.74 |
| PANSS pos | 38 (9/29) | 18.22 | 4.87 | 14.97 | 4.48 | -1.79 | 12.52 | 0.10 |
| PANSS neg | 38 (9/29) | 12.67 | 4 | 15.83 | 5.46 | 174.5* | - | 0.13 |
| PANSS gen | 38 (9/29) | 32.89 | 7.22 | 31.28 | 5.92 | -0.61 | 11.54 | 0.55 |
| Cognition |  |  |  |  |  |  |  |  |
| TMT A (sec) | 32 (9/23) | 30.33 | 11.95 | 34.48 | 14.36 | 121.5* | - | 0.46 |
| TMT B (sec) | 32 (9/23) | 75.00 | 43.81 | 99.74 | 70.39 | 132 |  | 0.24 |
| Inflammation |  |  |  |  |  |  |  |  |
| NLR | 41 (11/30) | 2.50 | 1.47 | 2.21 | 1.06 | 154* | - | 0.76 |
| MLR | 41 (11/30) | 0.32 | 0.13 | 0.27 | 0.09 | 133* | - | 0.36 |
| CSF |  |  |  |  |  | Chi2 |  |  |
| Q<sub>Alb</sub> evelation (y : n) | 25 (11/14) | 4:7 |  | 8:6 |  | 0.40 | 1 | 0.53 |

### Pharmacokinetic model analysis of DCE-MRI and K_trans_ maps

The open-source software ROCKETSHIP v. 1.2, 2016 [47] running with Matlab was used for DCE-MRI quantification and K_trans_ maps were calculated using Patlak modelling [48]. Quantitative pharmacokinetic modelling was chosen because it is easier to interpret and less sensitive to the acquisition protocol than qualitative and semiquantitative analyses, facilitating comparability between studies and sites [49]. The Patlak method was selected due to the low expected permeability and hence low likelihood of back-diffusion (transport of contrast agent from brain tissue back to blood) that are prerequisites for the use of the Patlak method [37]. Data processing in ROCKETSHIP included the following steps: Preparation of the dynamic datasets for DCE-MRI analysis, T1 mapping, selection of the AIF or reference region (transversal sinus), noise filtering. The fitted AIF from the individual datasets were used for the derivation of DCE-MRI parametric maps. (for more details see **suppl. methods**).

### DCE-MRI group comparisons

Group mean K_trans_ maps for the SSD and HC cohort are shown in the supplementary (**suppl. figures S3, S4**). To compare the whole-brain leakage (K_trans_ parameter maps) of cohorts, voxel-wise multiple regression or voxel-wise analysis of covariance (ANCOVA) were performed, while including covariates age, sex, BMI, smoking and education in the analysis. Compared groups were: SSD vs. HC (**hypothesis 1**), FEP (SSD) vs. HC and MEP (SSD) vs. HC (**hypothesis 2**). We then conducted linear regression analyses to investigate the leakage within the SSD group (K_trans_) in relation to psychopathology (PANSS total/positive/negative/general), cognition (TMT A, B), peripheral inflammatory markers (NLR, MLR)) (**hypothesis 3**) and BCSFB pathology/breakdown (Q_Alb_) (**hypothesis 4)**, correcting for age, sex, BMI, smoking and education. Significance threshold was set to voxel-wise alpha threshold of 0.001 and FWE cluster correction at an alpha of 0.05. All analyses were conducted in SPM12.

## Results

### Demographic data

A total of 45 individuals with SSD (11 females) and 42 HC (11 females) were included in the study (signed IC). Four individuals with SSD and two HC were declared as dropouts and excluded from the analyses (reasons for dropout are explained in **suppl. methods**), resulting in 41 individuals with SSD (10 females, mean age: 35.17 ± 9.77) and 40 HC (11 females, mean age: 33.55 ± 9.60) included in the final analyses. The SSD cohort was comprised by 35 individuals with schizophrenia, 5 with schizoaffective disorder, 1 with brief psychotic disorder. The average PANSS score was 62.47 (± 13.00, range 41-92), mean GAF 49.78 ± 12.51 (reflecting serious symptoms), 11 were in their first episode (FEP), 30 had multiple episodes (MEP), all were treated with antipsychotics, 15 had a lifetime history of clozapine intake (reflecting treatment resistance (TRS)). Mean duration of illness (DUI) was 9.2 years. 25 individuals had undergone CSF examinations; 12 out of the 25 showed an age-adjusted elevated Q_Alb_. Mean NLR was 2.29 ± 1.17 and MLR 0.28 ± 0.10.

Statistically significant group differences were detected in demographic aspects: total education (mean SSD= 14.84, mean HC= 18.39, p= 2.57e-06), BMI (mean SSD= 27.14, mean HC= 24.21, p= 0.0029)and cognitive assessments: TMT A (mean SSD= 33.31, mean HC= 23.10, p= 0.0009) and TMT B (mean SSD= 92.78, mean HC= 54.55, p= 0.0004). Sex and age did not differ significantly between the groups. **Table 1** demonstrates demographic data of the two groups and **table 2 and 3** the SSD cohort characteristics.

**Table 2.** SSD group, disease characteristics. Y= yes, n=no, TRS= treatment resistance, assessed through lifetime Clozapine intake, lifetime AP= lifetime antipsychotic treatment, CPZ= Chlorpromazine, GAF= Global Assessment of Functioning Scale PANSS= positive and Negative Syndrome Scale, pos= positive, neg=negative, gen= general, TMT= Trail Making Test

|  | **N** | **Mean** | **SD** |
| --- | --- | --- | --- |
| **Disease characteristics** |  |  |  |
| Diagnosis (DSM-5) | 41 | - | - |
| Schizophrenia | 35 | - | - |
| Schizoaffective disorder | 5 | - | - |
| Brief psychotic disorder | 1 | - | - |
| Duration of illness (DUI) (months) | 41 | 109.96 | 115.91 |
| First episode (y : n) | 11:30 | - | - |
| TRS (y : n) | 15:26 | - | - |
| Andreasen remission criteria (y : n) | 6:31 | - | - |
| CPZ equivalent [mg] | 41 | 406.84 | 271.41 |
| Lifetime AP (months) | 41 | 87.57 | 110.11 |
| GAF | 41 | 49.78 | 12.51 |
| **Psychopathology** |  |  |  |
| PANSS tot | 38 | 62.47 | 13.00 |
| PANSS pos | 38 | 15.74 | 4.72 |
| PANSS neg | 38 | 15.08 | 5.28 |
| PANSS gen | 38 | 31.66 | 6.18 |
| **Cognition** |  |  |  |
| TMT A (sec) | 32 | 33.31 | 13.67 |
| TMT B (sec) | 32 | 92.78 | 64.35 |

### DCE-MRI (K_trans_) group comparisons (hypothesis 1)

Whole K_trans_ mean maps were calculated for both groups (SSD, HC) (**supplemental Figures S3, S4**). The K_trans_ map comparisons of the two cohorts (SSD vs. HC) at a whole brain level showed significantly higher K_trans_ signal (leakage) in the SSD cohort compared to HC (**hypothesis 1**), when corrected for age, sex, BMI, smoking, total education. **Figure 1** illustrates the SSD/HC K_trans_ group comparison (main hypothesis). No brain region showed reduced leakage in the SSD cohort.

**Figure 1.**
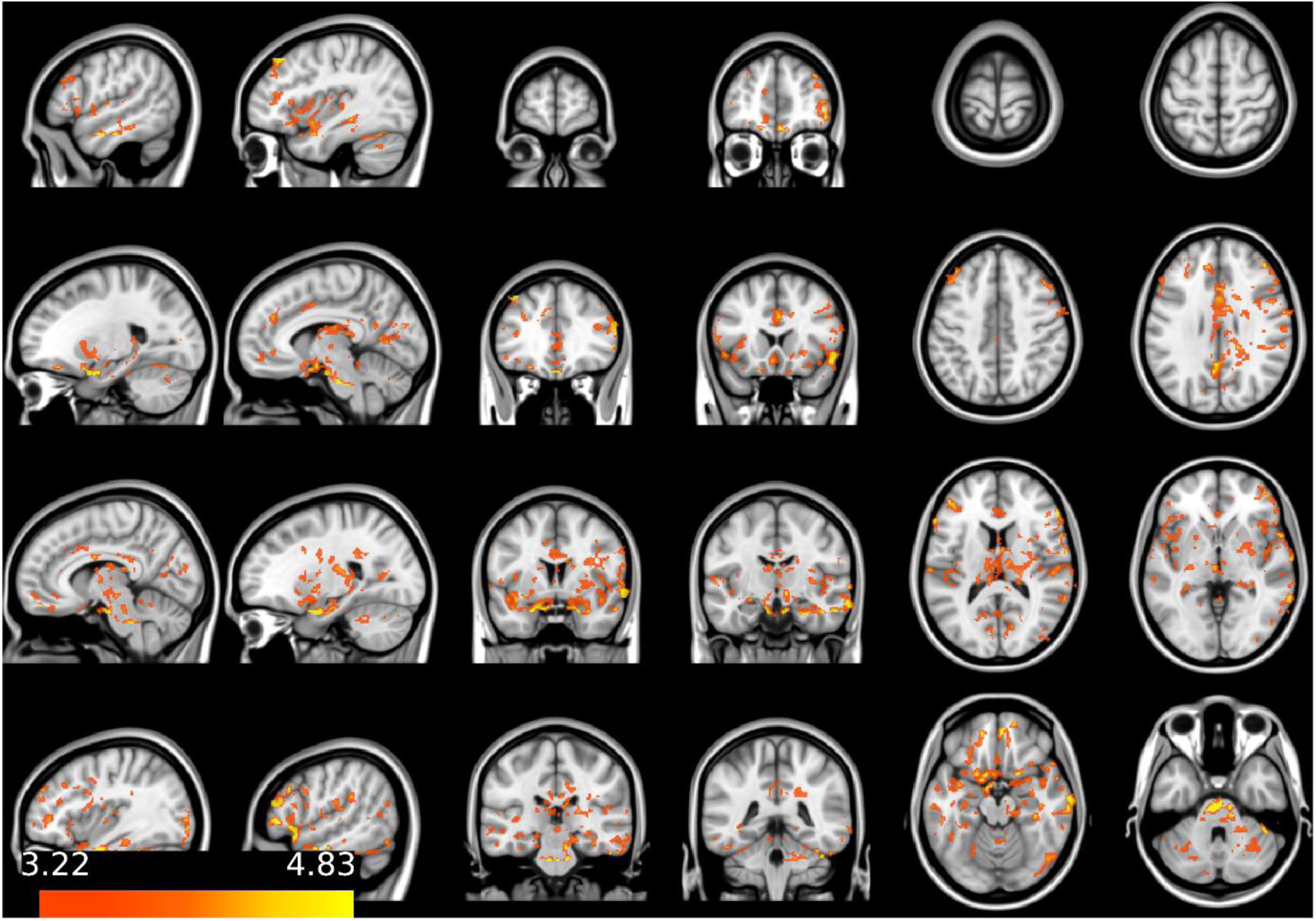
K_trans_ map comparisons of the two cohorts (SSD vs. HC) (SPM12) Higher K_trans_ (leakage) is detected in SSD patients, compared to healthy controls, color bar represents t-values

**Figure 2.**
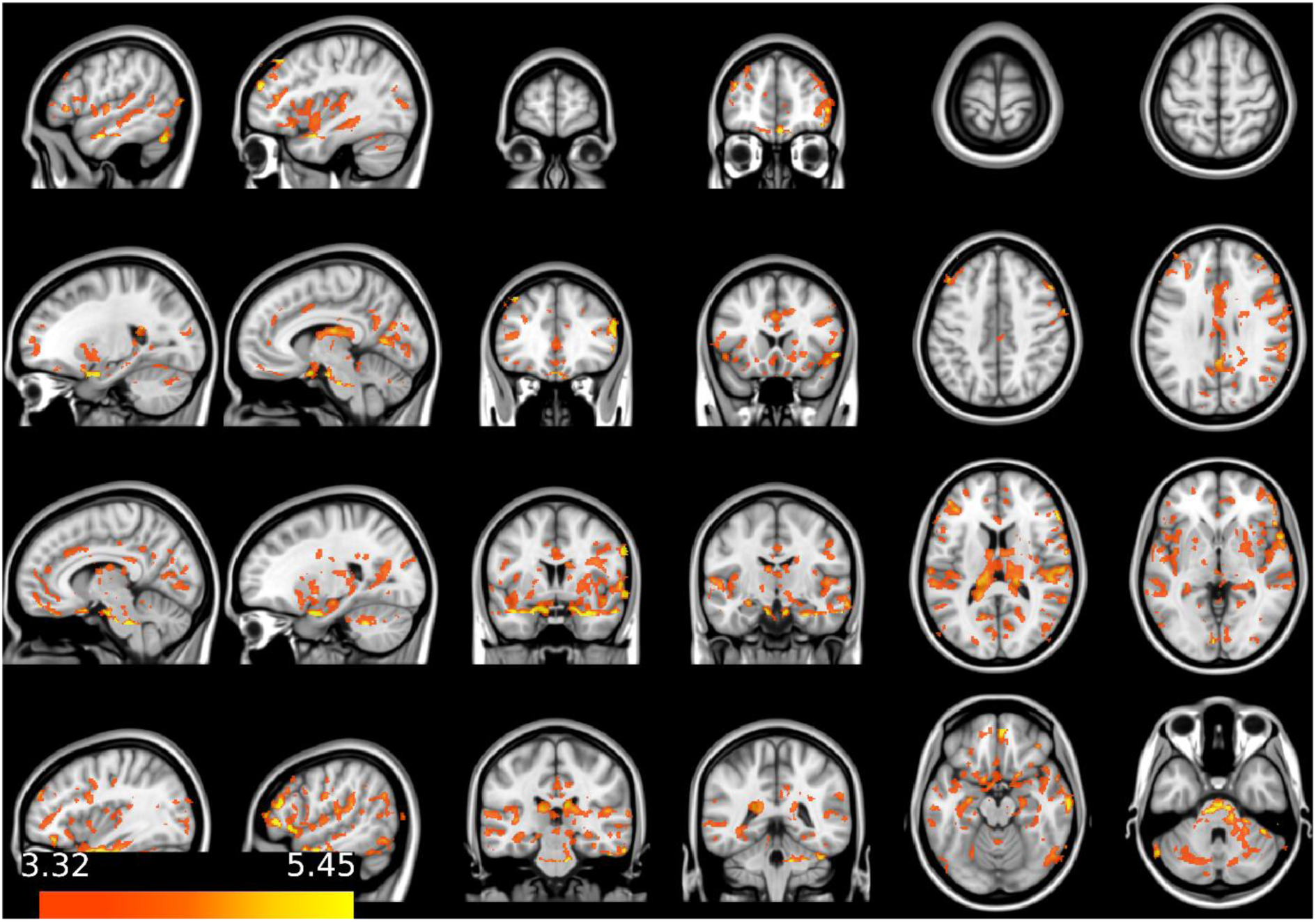
K_trans_ map comparisons of first-episode SSD (FEP) vs. HC (SPM12) Higher K_trans_ (leakage) in first-episode SSD patients, compared to healthy controls, color bar represents t-values

**Figure 3.**
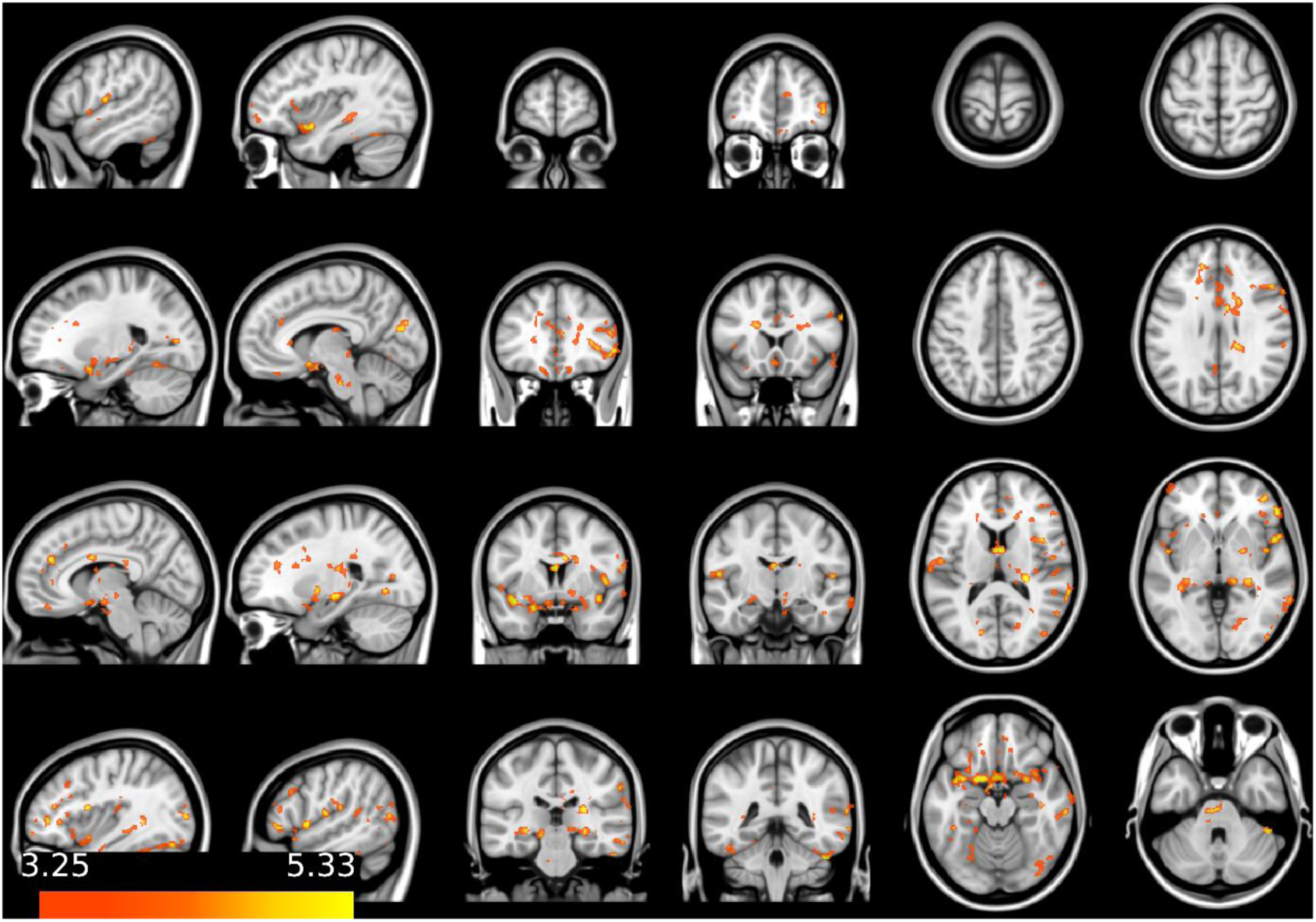
K_trans_ map comparisons of non-first-episode SSD (MEP) vs. HC (SPM12) Higher K_trans_ (leakage) in non-first-episode SSD patients, compared to healthy controls, color bar represents t-values

### Within group (SSD) tests: disease characteristics (hypothesis 2)

ANCOVA-based group comparisons, investigating the association of higher leakage (K_trans_) and stage of the disease (FEP vs. HC, MEP vs. HC) and linear regression models, investigating the association of higher leakage (K_trans_) and psychopathology/cognition (PANSS total, positive, negative, general, TMT A and TMT B), corrected for age, sex, BMI, smoking, total education, were performed. The analyses FEP vs. HC, MEP vs. HC showed a higher K_trans_ signal (leakage) in the FEP group compared to the MEP. **Table 3** shows demographic characteristics of FEP/MEP. Figure 2 illustrates the FEP vs. HC and Figure 3 the MEP vs. HC K_trans_ group comparison (**suppl. figure S5** shows the comparison of the FEP/MEP vs. HC effects).

Linear regressions testing for association of K_trans_ and psychopathology (PANSS total/positive/negative/general scores) and cognition (TMT A, B scores) showed no significant associations.

### Within group (SSD) tests: systemic inflammation and CSF (hypotheses 3, 4)

Linear regressions investigating the association of higher leakage (K_trans_) and peripheral inflammatory markers (NLR, MLR) (**hypothesis 3**), showed no significant results. Linear regression investigating the association of higher leakage (K_trans_) and Q_Alb_ (**hypothesis 4**), showed no association between higher K_trans_ values and elevated Q_Alb_.

## Discussion

The results of this study showed significantly higher K_trans_ values (representing BBB leakage) on a whole brain level in SSD, compared to HC, confirming our **main hypothesis** of higher BBB leakage in SSD. This is to our knowledge the second study so far investigating blood-brain barrier (BBB) leakage in individuals with SSD compared to HC with DCE-MRI. The only previous study investigating DCE-MRI in SSD with N= 29 SCZ vs. 18 HC reported higher leakage in SSD only in the thalamus [50]. The present data could replicate leakage in the thalamus (**suppl. fig. S6**). The additional effect localizations in the present study can be due to the significantly smaller sample size in Cheng et al. (total N= 47 vs. 81 in our study) and differences in cohort characteristics (Cheng et al. included younger subjects, almost half the cohort was untreated, and DUI was significantly lower with mean DUI= 42.6 vs. 109.96 months). Another reason for the difference in leakage localizations could be due to methodological differences. The scan time in Cheng et al. was 10 minutes including structural scans (vs. 22 minutes only DCE-MRI in the present study). A sufficient duration of the DCE-MRI sequence has been reported to be crucial when investigating very subtle barrier breakdown [27] and longer scan times improve the reproducibility [32]. The overall effect of higher leakage in SSD raises pertinent questions about the role of neurovascular dysfunction in the pathophysiology of this complex and heterogeneous disorder.

### Possible biological mechanisms leading to BBB impairment

Current evidence on the genetic and neurobiological underpinnings of SSD include neurodevelopmental and synaptic plasticity abnormalities, dysfunction in the neurotransmitter, microglial and oligodendrocytes system [51], and inflammatory pathways [52]. A growing body of evidence indicates abnormalities in vascular system [53, 54] and microcircuitry [51] in SSD, directly influencing the neurovascular unit (NVU), the minimal functional unit of the brain. Multiple brain regions have been shown to differ with respect to regional cerebral blood flow (rCBF) in patients (e.g. displaying frontal, temporal, parietal, cingulate and thalamic hypoperfusion [51, 55]), compared to healthy subjects. On a smaller scale, capillary abnormalities (such as thickening, deformation, vacuolation of basal lamina, prominent swelling and vacuolation of astrocytic end-feet, alterations of pericapillary oligodendrocytes and signs of activation of microglial cells) have been described post-mortem and are believed to occur during brain development and to continue in the course of illness in SSD patients [56]. Also, significantly reduced claudin-5 levels, a tight junction protein in SSD-relevant brain regions such as the hippocampal grey matter, and altered expression of junctional components, have been reported [57]. These changes in the microenvironment of the NVU directly influencing the BBB (an NVU component) have been postulated to be induced by multiple mechanisms like chronic hypoperfusion, metabolic disturbances leading to a disrupted cellular osmoregulation and inflammation [56], and are the structural underpinnings possibly explaining a higher leakage in the SSD cohort. These changes have in turn been reported to possibly lead to hyperpermeability, neuroinflammation and oxidative stress [3, 58].

### Parallels with neurodegenerative disorders

The disruption in the tightly regulated relationship between neuronal activity and blood flow (neurovascular coupling) can further result in disturbed oxygen metabolism, neuronal death, and brain tissue atrophy [55]. Of note, the ultrastructural changes reported in SSD resemble changes in the aging brain [56]. The association of BBB disruption and neurodegeneration and associated cognitive decline has also frequently been described in neurodegenerative disorders (NDD) [8, 59]. This poses the question of related pathomechanisms between SSD and NDD, as postulated over a century ago under the term “dementia praecox” [60] and supports the theory of SSD being a cognitive disorder [61]. Interestingly, even though cognitive function (TMT A and B scores) differed significantly between SSD and HC in the present study, K_trans_ values did not correlate with cognitive dysfunction within the SSD cohort, challenging the association of leakage and cognitive decline. However, this can be due to the limited cognitive domains the TMT covers (mainly attention, visual scanning, mental flexibility, and executive functions), hence other cognitive functions (e.g. working memory, semantic memory, verbal fluency, could potentially correlate with K_trans_. Also, the within cohort regressions were possibly underpowered due to reduced sample size.

### The role of cardiovascular risk factors

When interpreting BBB disruption in the context of disturbed microvasculature, comorbidities frequently accompanying SSD, leading to peripheral vascular endothelial dysfunction and associated increased cell permeability [62], have to be discussed as possible contributors, such as metabolic syndrome, cardiovascular disease, and diabetes mellitus [63]. This association was also confirmed in our SSD cohort, that displayed significantly higher BMI than HC. Even though we controlled for the covariate BMI and smoking, the full extent of cardiovascular risk factors (CVDRF) and their impact on the BBB could not be fully eliminated in the statistical models. Of note, a recent meta-analysis reported global cognitive deficits in individuals with (vs. without) metabolic syndrome. Among the metabolic syndrome disorders, diabetes and hypertension were associated with significant cognitive impairment, while obesity was not [63]. This may be explained by varying degrees of influence that each CVDRF exerts on endothelial function and the BBB.

### Pathomechanisms in the context of disease course and inflammation

The etiology of the previously reported post-mortem ultrastructural changes that possibly underly BBB leakage has partly been attributed to chronic hypoperfusion leading to BBB damage [56]. This is due to the known alterations on a cellular level, associated with chronic cerebral hypoperfusion [64], suggesting a positive correlation with illness chronicity. However, this association could not be verified post-mortem [56] and in vivo studies also demonstrated no link between CBF abnormalities and disease chronicity [56, 65]. These data challenge the theory of BBB disrupting effect of disease chronicity and indicate these changes could be caused during neurodevelopmental stage, which is also supported by the established link of hypoxia and vascular factors as interacting elements in the neurodevelopmental model [53].

Of note, higher leakage (K_trans_) values in the present study were more prominent in individuals with first episode psychosis (FEP), indicating a sub-phenotype of SSD (**hypothesis 2**), and further contradicting the chronicity theory. FEP have been associated with reduced antioxidant status, a pro-inflammatory imbalance [66] and higher pro-inflammatory cytokine (PIC) levels [67], suggesting inflammatory mechanisms to be of high relevance in the initial disease stage [68]. This is of clinical relevance, as inflammatory processes such as high levels of PICs have been reported to impede early symptomatic remission [69]. Circulating PICs (e.g. TNF-alpha) have been reported to be involved in changes in astrocytic end-feet through the release of nitric oxide in the rat brain [70], directly linking (early) inflammatory processes with BBB disruption. It is possible that the difference in K_trans_ between FEP and MEP reflects an inflammatory reaction early in the disease course. Interestingly, in the present study, individuals with FEP presented higher mean values of peripheral inflammatory markers (NLR, MLR), than MEP. These have been reported to be elevated in SSD, and in FEP [71], compared to HC [39, 71, 72]. The difference in mean values was however not significant in our cohort and K_trans_ showed no association with MLR/NLR (**hypothesis 3**). This could be because these ratios only permit the evaluation of the *cellular* immune reaction and mostly allow estimations on *peripheral* rather than *CNS* inflammation, mainly due to their size. Cells physiologically transfer through the blood-meningeal barrier as it is more permeable than BBB [73]. They might hence not be the most reliable markers in the context of subtle neuroinflammation and BBB disruption. The lack of association underscores the difficulty to bridge the brain and the periphery, especially in the context of *subtle* inflammatory mechanisms.

Taking these data into account, it is possible, that the different contributing pathways in BBB disruption compose a "three-hit hypothesis", first hit being in neurodevelopmental stage, second hit being inflammatory reaction, with increased PICs, abnormal CBF and aggravation of BBB disturbance (FEP) and third hit being the chronic hypoperfusion combined with culminating cardiovascular risk factors, further damaging or maintaining the damage in the BBB (MEP). This theoretic model strengthens the genetic–inflammatory–vascular theory of SSD that intertwines neurodevelopmental, -degenerative and -inflammatory processes [74], rather than interpreting them as competing theories.

### Correlation with Clinical Symptoms

Cheng et al. reported significant positive correlations between thalamic mean K_trans_ values and symptom severity. This could not be replicated in this study. A possible reason for the difference in results could be the more severe psychopathology in the Cheng et al. cohort (mean PANSS 80.79 vs. 62.47 in our cohort). Of note, evidence on ultrastructural changes in vasculature in SSD also showed no differences between SSD clinical subgroups (negative/positive symptoms, paranoid/nonparanoid types, different course of paranoid schizophrenia) [56]. Further studies investigating the association of BBB leakage and psychopathology are needed.

### Correlation with Q_Alb_

The present study showed no association between K_trans_ and Q_Alb_ that represents BCSFB disruption (**hypothesis 4**). Interestingly, another recent study investigating DCE-MRI and Q_Alb_ in dementia also reported no correlation between Q_Alb_ and K_trans_ [75]. This further underscores the differences in structural and pathophysiological properties between BBB and BCSFB and the importance of differentiation between the two.

### Limitations and future directions

Our findings must be interpreted in the context of their limitations. Firstly, although we corrected our statistical models for BMI, we did not correct for all CVDRF possibly mediating BBB disruption. Further limitations involve a sampling bias, as not all patients eligible for CSF diagnostics were included in our monocentric study. Another factor possibly mediating BBB changes, is antipsychotic treatment (AP), that can influence barrier functionality [76]. Our cohort did not include medication-naive patients, however, the lower CPZ equivalents and shorter lifetime AP treatment duration in FEP, alongside higher leakage, suggest that AP is unlikely to be the primary cause for the increased leakage observed. Another limitation pertains to the method of pharmacokinetic models, that are based on a highly simplified description of tissue microstructure and function and can hence ignore potentially relevant features such as interstitial fluid transport [49]. However, the combination of DCE-MRI with the Patlak model has been described as the method of choice in permeability imaging with subtle expected leakage [37] and is recommended in the HARNESS (*HARmoNising Brain Imaging MEthodS for VaScular Contributions to Neurodegeneration* [77]) consensus recommendations [49]. While this is the study with the largest SSD cohort undergoing DCE-MRI compared to HC and the total number of N=81 is notably higher compared to available DCE-MRI studies [49], for the subgroup analyses our sample size remained relatively small. Lastly, the cross-sectional nature of the approach does not allow conclusions on dynamic changes in the BBB that possibly underly the disorder. Larger longitudinal, multimodal studies, that investigate the temporal relationship between BBB integrity and disease progression, combined with vascular and BBB genomics are essential to robustly replicate our findings and comprehend the relevance and dynamics of BBB disturbances in SSD.

## Conclusion

In conclusion, the present study is the first to demonstrate BBB leakage through DCE-MRI on a whole brain level in SSD compared to HC. The only previous study performing DCE-MRI in SSD included 29 SSD and 18 HC. The present study, almost doubling this sample size with N= 82 is the study with the largest cohort to undergo this investigation, in the context of a broader spectrum of assessments, including psychopathology, cognition, disease course data, blood derived immune-related and CSF markers. Finally, this is the first study to conduct DCE-MRI with sufficient duration of the sequence. The reported effects are possibly mediated by ultrastructural changes in the NVU reported in post-mortem studies and the result of a complex interaction of neurodevelopmental and -inflammatory processes in the context of the genetic–inflammatory–vascular theory of SSD. The findings confirm the hypothesis of a disrupted BBB in SSD and suggest that DCE-MRI is a useful tool in SSD diagnostics. Understanding the role of the BBB in SSD could open avenues for targeted interventions aimed at preserving or restoring BBB integrity. Moreover, the integration of DCE-MRI assessments of BBB integrity with existing diagnostic tools holds promise for enhancing early detection of biological mechanisms in SSD.

## Data availability

The de-identified data of this study will be made available in BIDS format upon publication in the Open Neuro repository (https://openneuro.org/) (link accessible after acceptance of the manuscript).

## Code availability

All software packages used in this study are publicly available:

Rocketship v1.2 2016 (https://github.com/petmri/ROCKETSHIP/blob/master/dce/compare_gui.m),

FSL v. 6.0.5.1 (https://fsl.fmrib.ox.ac.uk/fsl/fslwiki/FslInstallation ),

SPM12, v. 7771 (https://www.fil.ion.ucl.ac.uk/spm/software/spm12/)

## Author contributions

This research was not supported by any specific grant from funding agencies in the public, commercial, or not-for-profit sectors. JM was supported by the Förderprogramm für Forschung und Lehre (FöFoLe), University Hospital, LMU Munich (project number 1167).

EW, DK and JM designed and conceptualized the study with supervision of PF. EW, DK, JM designed this study and wrote the protocol. JM recruited patients and collected study data. DK and KS trained staff on MRI assessments. MRI acquisition were performed by JM, JuM, BP, IL. MRI preprocessing was performed by HT, IL. NF supervised and performed MRI data analysis. All statistical analyses were performed by NF and JM. Data visualization was performed by NF. JM wrote the first draft of the manuscript. All authors contributed to and approved the final version of the manuscript.

## Acknowledgments

The procurement of the MRI scanner was supported by the Deutsche Forschungsgemeinschaft (DFG, German Research Foundation) grant for major research (DFG, INST 409/193-1 FUGG). The authors thank all participants for their contribution. The authors thank Dr. Hanna Zimmermann and Dr. Thomas David Fischer for their help in clinical evaluation of the MRI scans.

