## Supplementary material for "Increased blood–brain barrier leakage in schizophrenia spectrum disorders compared to healthy controls in dynamic contrast-enhanced magnetic resonance imaging": suppl. information

### Supplemental information

#### Supplemental tables

##### Table S1

**Table S1**

**DCE-MRI methodology used for measuring BBB permeability**

| | |
|---|---|
| **T1 mapping** | |
| T1 mapping method used? | MPRAGE |
| **Contrast agent** | |
| Contrast agent used | Gadovist |
| CA dosage used | 0.1 mmol/kg |
| CA infusion rate | 3 ml/s |
| CA dosage varied? | No |
| **Acquisition factors** | |
| Scanner type | 3T/Siemens MAGNETOM Prisma |
| Baseline measurements | Yes |
| Sequence details | Voxel size 1.8 x 1.8 x 1.8 mm, TR 5.00 ms, TE 2.21 ms, flip angle 15.0 deg, FOV 173 x 173 x189 mm, slice thickness 1.8mm, CA injected after 2 min, total duration 23 min |
| **tracer-kinetic model** | |
| **Model selected** | Patlak |

TE= echo time, TR= repetition time, FOV= field of view, CA= contrast agent

### Supplemental figures

**Figure S1**

**CSF circulation**

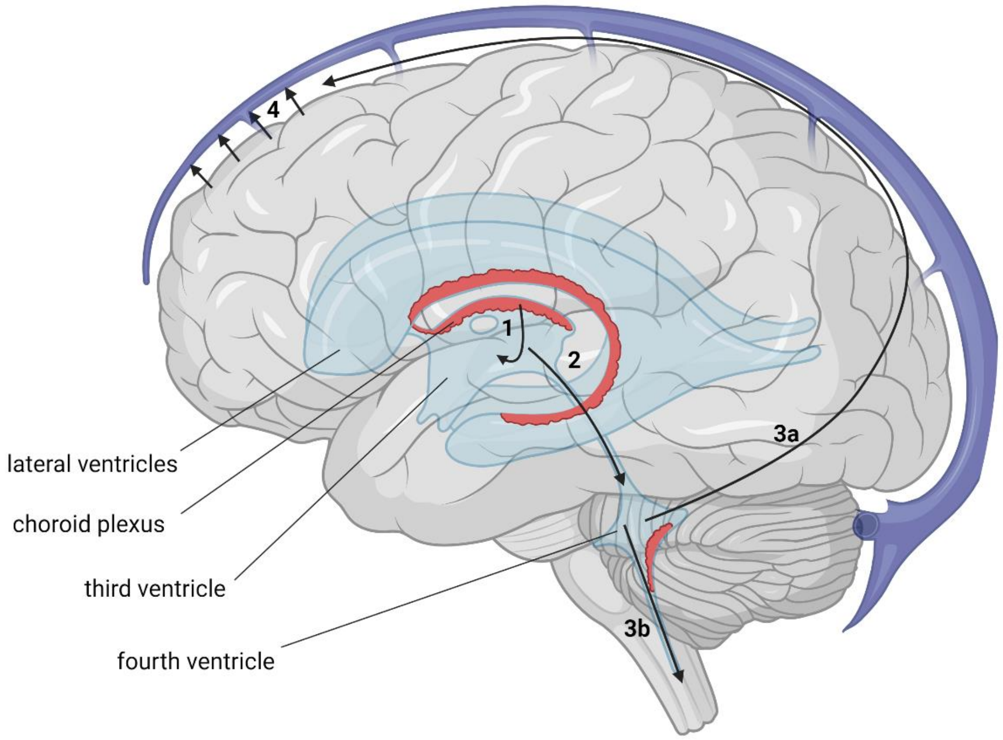


The CSF is produced by the choroid plexus and moves from the lateral, third and fourth ventricle, either upwards across the surface of the brain and through lymphatics and the arachnoid granulations back into the blood stream, or downwards into the spinal canal, limiting the direct relevance of $Q_{Alb}$ in the context of BBB evaluation. Created with BioRender.com

### Figure S2

**Figure S2**

**DCE-MRI data acquisition and processing, Created with BioRender.com**

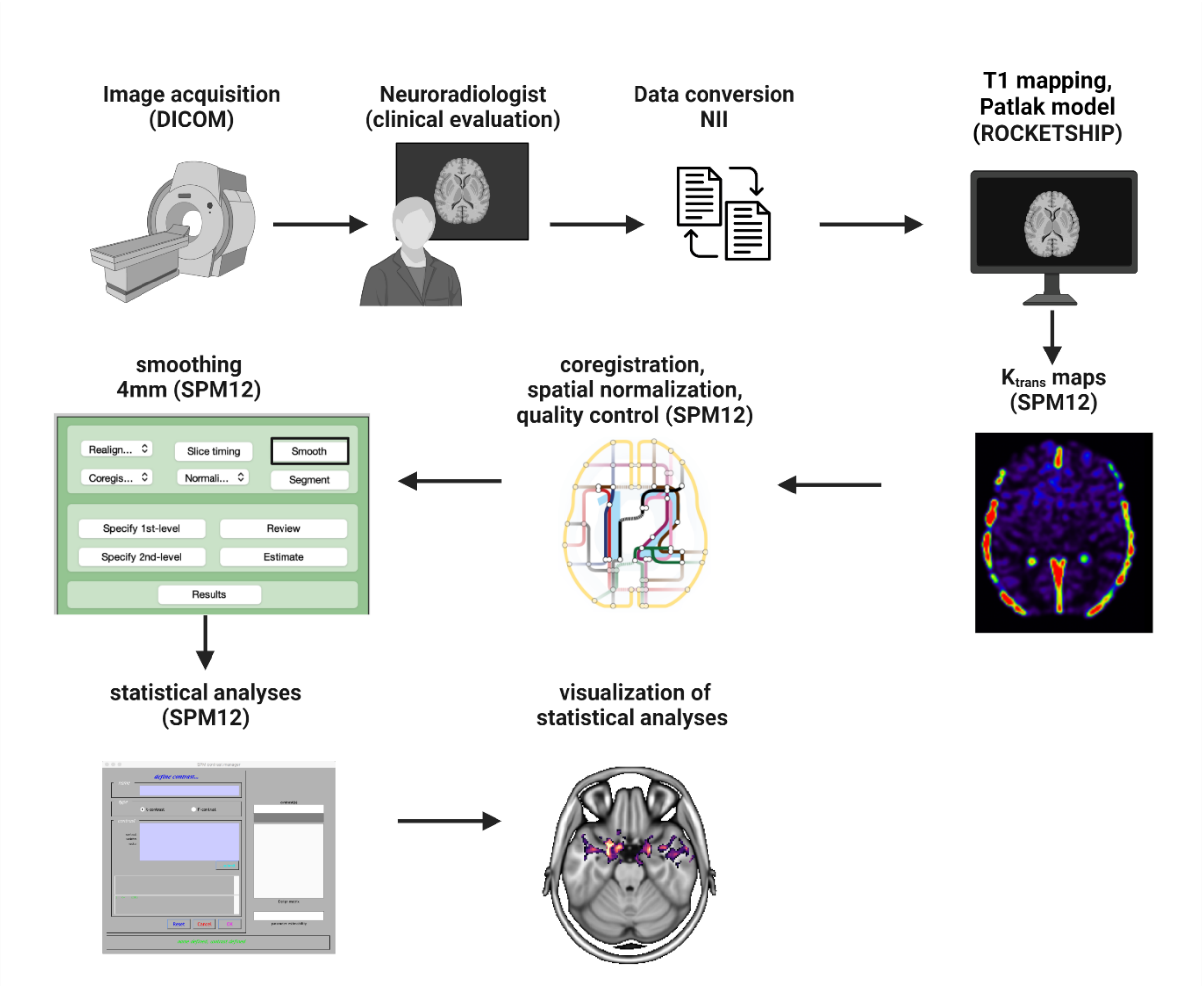

### Figure S3

**Figure S3**

**$K_{trans}$ means SSD cohort (SPM12)**

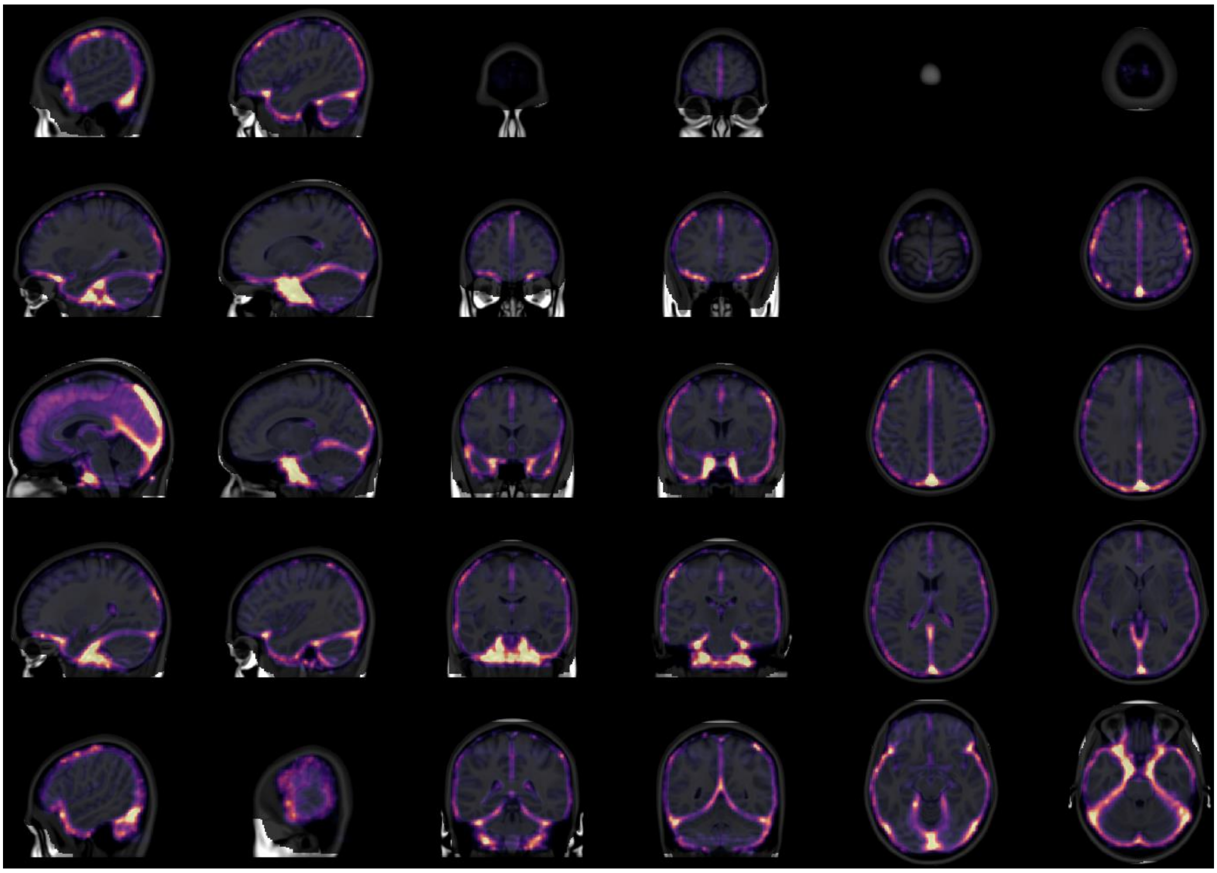

### Figure S4

**Figure S4**

**$K_{trans}$ means HC cohort (SPM12)**

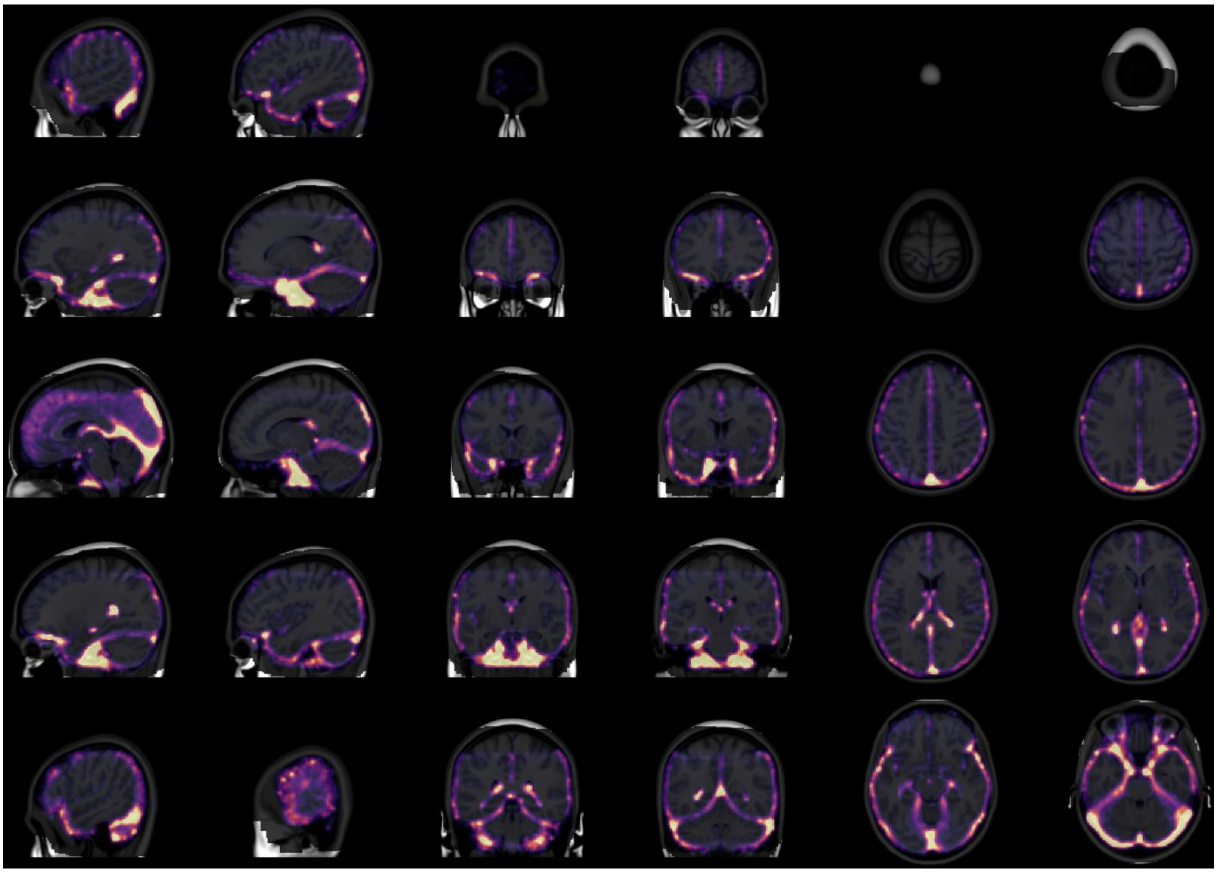

### Figure S5

**Figure S5**

**FEP vs. HC and MEP vs. HC (SPM12)**

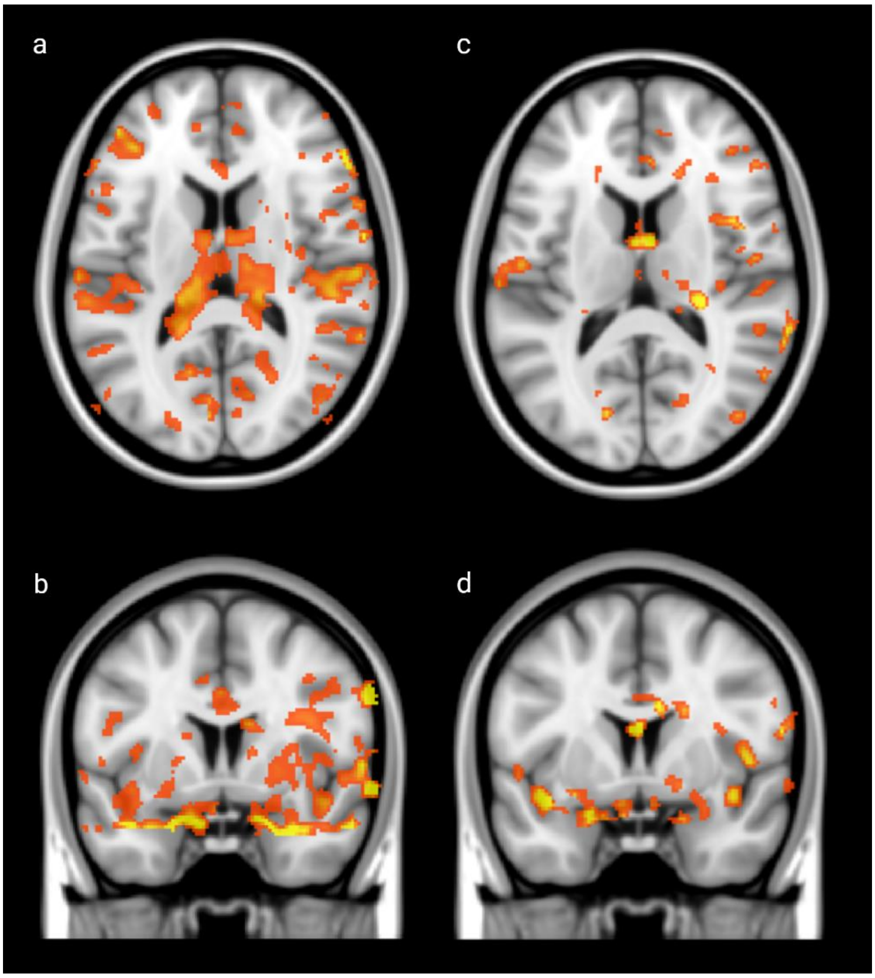


The effect of higher $K_{trans}$ (leakage) in SSD patients compared to HC is stronger in first-episode SSD (a, b) than in multiple episode SSD (c, d)

### Figure S6

**Figure S6**

**$K_{trans}$ map comparisons of first-episode SSD (FEP) vs. HC (SPM12) showing higher leakage in thalamus (a), Created with BioRender.com**

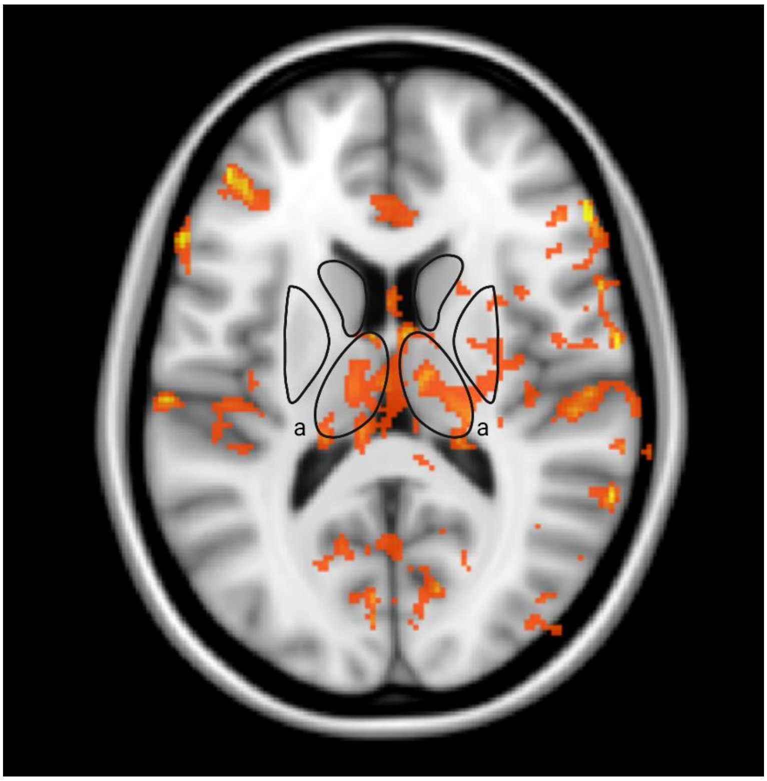

### Supplemental methods

#### Dropouts

Four individuals with SSD were excluded from the DCE-MRI analyses due to structural abnormality in the MRI (SSD-026), premature termination of the scan by the individual (SSD-021) and technical issues (SSD-029, SSD-077). Out of the heathy control group two individuals were excluded from the analyses due to an autoimmune (HC-017) and psychiatric condition (HC-018).

#### CSF autoantibodies

The CSF samples [total n = 25], were analyzed in the laboratory of our hospital by a cell-based assay from EUROIMMUN (Lübeck, Germany) for autoimmune encephalitis (antibodies tested: NMDAR [n = 25], α-amino-3-hydroxy-5-methyl-4-isoxazolepropionic acid receptor-1 (AMPAR-1) [n = 25], α-amino-3-hydroxy-5-methyl-4-isoxazolepropionic acid receptor-2 (AMPAR-2) ([n = 25], CASPR2 [n = 25], LGI1 [n = 25] and γ-amino-butyric acid receptor B1/B2 [n = 25]). No antibodies were detected in CSF and only one subject (SSD-047) manifested NMDAR antibodies in serum.

#### Quantification of the Blood-Brain Barrier Permeability

DCE-MRI analysis was performed using Rocketship software (version 1.2 2016) running with Matlab. To account for a possible confounding effect of blood flow on DCE-MRI measurements, we determined the arterial input function (AIF) curve for every individual, to provide a dynamic profile of GBCA concentration in the blood after the i.v. injection [1]. Due to the easy assessment (size, location and easier accessibility than for example internal carotid artery), the concentration of the tracer in plasma (Cpa(t)) was determined from the time course of the signal change in a pixel in the transverse sinus (TS), assuming proportionality to the arterial concentration of the GBCA [2]. Using the individual AIF dynamic profile measurements of the GBCA concentration in the blood self-corrects for possible differences in the blood flow that may affect delivery of the CA to the brain and could potentially affect the $K_{trans}$ measurements [1]. The AIF, which was extracted from a region-of-interest (ROI) positioned at the TS, was fitted with a bi-exponential function prior to fitting with the Patlak model [3]. The Patlak linearized regression mathematical analysis was used to generate the BBB permeability $K_{trans}$ maps. This model requires that the CA diffusion across the BBB remains unidirectional during the acquisition time. The total tracer concentration in the tissue, $C_{tissue}$ (t), can be described as a function of the blood concentration, $C_{AIF}$ (t), the intravascular blood volume, $v_p$, and a blood-to-brain transfer constant, $K_{trans}$, that represents the flow from the intravascular to the extravascular extracellular space using the equation:

$$C_{tissue}(t) = K_{trans} \int_0^t C_{AIF}(u)\, du \; + \; v_p C_{AIF}(t)$$ [1, 4]
